# Gestational diabetes mellitus and vascular malperfusion lesions in the placenta: a systematic review and meta-analysis

**DOI:** 10.1101/2025.01.29.25319856

**Authors:** Amrita Arcot, Rachel E. Walker, Kelly Gallagher, Jeffery A. Goldstein, Alison D. Gernand

**Affiliations:** The Pennsylvania State University Department of Nutritional Sciences, University Park, PA 16802; The Pennsylvania State University Ross and Carol Nese College of Nursing, University Park, PA 16802; Northwestern University Feinberg School of Medicine Department of Pathology, Chicago, Illinois 60611

**Keywords:** Gestational diabetes mellitus, GDM, placenta, placental lesions

## Abstract

**Background:** Gestational diabetes mellitus (GDM) can result in increased placental lesions related to high maternal blood glucose, but these relationships are not well understood.

**Objective:** To examine the relationship between GDM and placental vascular malperfusion lesions: accelerated villous maturation, increased syncytial knots, delayed villous maturation, and increased fibrin deposition.

**Search strategy:** PubMed, BIOSIS, and Web of Science databases were systematically searched for full-text articles in English from inception until August 21, 2024.

**Selection criteria:** Our inclusion criteria were randomized controlled trials, case-control, cohort, and cross-sectional studies that examined the relationship between GDM and selected placental vascular malperfusion lesions. The outcome must have been reported as a total proportion.

**Data collection and analysis:** We included all eligible studies in narrative synthesis. If an outcome of interest was in at least three studies, we calculated the odds ratios (OR) by GDM diagnosis, with 95% confidence intervals, using mixed-effects logistic regression with random study effects. We evaluated the risk of bias with the Newcastle-Ottawa Scale.

**Main results:** We screened 151 studies, of which eight were included (n=1,291), and six met the criteria for meta-analysis (n=561). Unadjusted odds (95% confidence interval) of delayed villous maturation were six-fold higher (OR: 6.37 [3.28, 12.37]) in pregnancies with GDM compared to those without GDM. The narrative synthesis of the literature found higher proportions of increased syncytial knots, delayed villous maturation, and increased fibrin deposition, but not accelerated villous maturation, in pregnancies with versus without GDM.

**Conclusions:** GDM was associated with increased risk of three placental malperfusion lesions, although there is a small number of studies in this area. Future investigations should examine if these vascular malperfusions are associated with adverse pregnancy outcomes often linked with GDM.

**Synopsis:** GDM is associated with increased syncytial knots, delayed villous maturation, and increased fibrin deposition, but not accelerated villous maturation.

**Funding:** Health Resources and Services Administration (HRSA) of the U.S. Department of Health and Human Services (HHS) as part of the Maternal Child Health Bureau Nutrition Training Grant, The TRANSCEND Program in Maternal Child Health Nutrition (T7949101).

**Registration:** Registered in PROSPERO under protocol number CRD42023449651

## Introduction

Gestational diabetes mellitus (GDM) is defined by glucose intolerance with first recognition during pregnancy and no previous history of type 1 or type 2 diabetes mellitus.^1^ In 2021, 8.3% of live births in the United States presented with GDM (approximately 305,000 births).^2^ That same year the global rate of hyperglycemia in pregnancy was 16.7%, of which 80.3% (16.9 million) was attributed to GDM.^2^ Previous research shows that GDM leads to an increased risk of neonatal respiratory distress, delivering a large-for-gestational age infant, Cesarean section, admission to an neonatal intensive care unit, and other adverse pregnancy outcomes.^3,4^ Some studies have found that pregnant persons with GDM also have altered placental characteristics, such as increased placental weight^5^ and placental lesions on the maternal and fetal side.^6,7^ Placental abnormalities can have a downstream impact on neonatal outcomes (e.g., stillbirth, neonate ventilation, etc.).^8^

The placenta is a complex, highly vascularized organ that supports fetal growth and development, and it produces hormones to facilitate nutrient signaling between the maternal and fetal circulation.^9^ It functions to transport oxygen and nutrients to the fetus and flow oxygen-depleted blood and waste from the fetus back to the maternal body. Past evidence links GDM with both maternal and fetal vascular malperfusion, which includes accelerated villous maturation (AVM), increased syncytial knots (a characteristic of AVM), delayed villous maturation (DVM), and increased fibrin deposition.^6,10–12^ Other complications include villous infarcts, fibrinoid necrosis, villous edema, and other complications.

To our knowledge, published reviews of GDM and the placenta have been narrative in nature and broadly examined GDM and placental characteristics, encompassing placental morphology and histopathological outcomes.^6,7^ Therefore, we aimed to conduct a systematic review and meta-analysis to examine the relationship between gestational diabetes mellitus and vascular malperfusion lesions in the placenta. We selected four common vascular lesions to examine: two on the maternal side (AVM and increased syncytial knots) and two on the fetal side (DVM and increased fibrin deposition).

## Methods

### Information sources

We conducted a systematic review and meta-analysis of original, peer-reviewed studies reporting an association between GDM and placental vascular malperfusion lesions. The outcomes of interest were based on frequently identified placental lesions in pregnancies with diabetes.^6,7^ The review protocol was preregistered in PROSPERO on August 08, 2023 (PROSPERO ID #CRD42023449651). This systematic review followed the Preferred Reporting Items for Systematic Reviews and Meta-Analyses (PRISMA) guidelines.^13^ PubMed, BIOSIS, and Web of Science databases were systematically searched from inception until August 2, 2023, using search terms for GDM, AVM, increased syncytial knots, DVM, and increased fibrin deposition (**Table S1**). Additionally, we searched for studies by reviewing the reference list of published papers. An updated search was conducted on August 21, 2024, to account for any new publications since the original search.

We selected two common lesions for maternal vascular malperfusion (AVM and increased syncytial knots) and fetal vascular malperfusion (DVM and increased fibrin deposition).^10–12^ Of note, increased syncytial knots are a characteristic of AVM.^14^

### Eligibility criteria

We included studies in this systematic review if they met the following criteria: (1) study design: randomized controlled trial, case-control, cohort, or cross-sectional; (2) exposure: studies that specified GDM; (3) exposure: study explicitly stated or adequately described GDM criteria (**Table S2**); (4) outcome: studies that specified placental AVM, syncytial knots, DVM, and/or fibrin deposition; (5) outcome: measured as a total proportion (n, %) of placenta samples or reported in a way that total proportion was calculable for GDM and comparison groups separately; (6) comparison of GDM to a healthy, normal, broad, or general obstetric population. We excluded articles if they were conducted in non-English language or were case reports, non-human studies, unpublished studies, or not peer-reviewed. The Cochrane Collaboration requires a minimum of two similar studies to conduct a meta-analysis;^21^ we were conservative and required at least three. Six studies reported DVM and therefore qualified for meta-analysis. Four studies examined fibrin deposition,^15–18^ but tissues were sampled from different placental locations, thus results were incompatible with meta-analysis. Three studies examined syncytial knots, but one reported the total proportional mean and standard deviation of syncytial knots by GDM groups (as opposed to the proportion of *increased* syncytial knots). As such, increased syncytial knots were not eligible for meta-analysis.

### Study selection

Two reviewers (AA and KG) independently screened titles and abstracts and were blinded to the other reviewer’s answers so that they could evaluate each search result twice. Once the first review was complete, AA unblinded all decisions to determine the number of articles that remained *undecided* and in *conflict*. *Undecided* studies underwent a full-text review by both reviewers to assess their inclusion or exclusion in the abstraction stage. In cases of conflict, the two reviewers discussed each study until a consensus was met. JAG and KG were our clinical experts for all stages of the review. Rayyan’s web application was used to manage the title/abstract screening stage.^19^ One reviewer (AA) screened all titles and abstracts in the updated search.

### Data extraction

In the full-text abstraction stage, two reviewers (AA and REW) assessed eligibility (see *inclusion and exclusion*) and independently abstracted data from all studies that passed the title and abstract review in the original search. Any disagreement was discussed between both reviewers until a consensus was met. The updated search yielded four additional studies, and one reviewer (AA) conducted the full-text abstraction. A Microsoft Excel sheet was used to collect the following information from all studies: citation details, study type, location (city, country), years the study was conducted, sample size, participant characteristics, delivery data (e.g., gestational age, birth weight, and placenta weight), GDM diagnostic criteria, placental preparation and timing of placental preparation, and main findings as reported by each study. We also collected proportion details (n, %), and/or data that could calculate a proportion, for all four placental lesion outcomes of interest by GDM and comparison group. Following data abstraction, one reviewer (AA) compared all information to the original study and checked for discrepancies. Any discrepancies in abstracted data were discussed until both reviewers reached a consensus.

### Assessment of risk of bias

We assessed the risk of bias in all eligible studies using the Newcastle Ottawa Scale for case-control studies.^20^ A higher total score indicates a lower risk of bias. Both reviewers (AA and REW) independently assessed all studies in the initial search, followed by a discussion to identify and resolve any disagreement. The quality of studies was categorized based on past literature, which utilized the Newcastle Ottawa Scale: 0-2 (poor), 3-5 (fair), 6-9 (good).^21^ One reviewer (AA) conducted the risk of bias for the one study eligible for systematic review after the updated search.

### Data synthesis without meta-analysis

We first summarized all studies in the systematic review, regardless of inclusion in the meta-analysis.^22^ Studies not eligible for meta-analysis were described using narrative synthesis by grouping and reporting by outcome of interest (AVM, increased syncytial knots, and increased fibrin deposition).^22^ Individual participant data was not present or retrieved for any study; therefore, all results were described by their total proportion (n, %). Effect sizes were reported if present in study results. Finally, results across studies are summarized by lesion.

We combined studies for narrative synthesis and meta-analysis for tabular presentation of methods and study characteristics. Studies were listed in reverse chronological order. Next, we compiled proportion (n, %) of lesions and statistical differences in lesions for GDM vs non-GDM controls reported by the original study for each lesion in our study aim. Additionally, we reported the same data on lesions that were outside of the aim of the study. Then we synthesized findings of (1) placental weight (mean, standard deviation) and (2) placental efficiency (that is placental:fetal weight ratio) by GDM group due to the past literature on the association between GDM and placental morphology.^7^

### Data synthesis and meta-analysis

We used mixed-effects logistic regression with random study effects to estimate the odds of DVM if diagnosed with GDM.^23^ Collected data were unadjusted, as a result all calculated point estimates were unadjusted with 95% confidence intervals. *Schäfer-Graf et al.* reported DVM in three categories: normal/mild (no distinction between normal and mild), moderate, and high.^23^ We categorized normal/mild as *normal* in the meta-analysis (non-GDM, n=58; GDM, n=66).

Heterogeneity between studies was evaluated using I^2^ and Cochrane’s Q test.^25^ The Cochrane Collaboration interprets an I^2^ of 0-40% as low, 30-60% as moderate, 50-90% as substantial, and ≥75% as considerable heterogeneity.^26^ Publication bias was visually inspected by the symmetry of effect sizes in a contour-enhanced funnel plot.^27^ We did not use formal statistical analysis, like Egger’s test, because it is discouraged with small study samples (k ≤ 10) due to lack of test power.^28,29^

Sensitivity analysis was conducted using the *leave-one-out method*.^6^ In this method, one study is removed in each cycle, and the pooled unadjusted odds ratio is re-estimated using mixed-effects logistic regression models.^26^ Influence analysis plots illustrated multiple diagnostic tests to check the quality of regression fit for each study.^30–33^ Studies that contribute the most to the variability in effect sizes were excluded, and heterogeneity was re-analyzed. Methods to examine heterogeneity (e.g., subgroup analysis and meta-regression) are discouraged with small study samples (k ≤ 10) and were thus not included in our analysis.^34^

We conducted all analyses in R, version 2024.04.2.^35^

## Results

### Study selection and study characteristics

**Figure 1** describes the study selection process. We assessed 30 studies for eligibility in the full-text abstraction. Eight met the criteria for this systematic review,^15–17,24,36–38^ of which six were eligible for meta-analysis.^15,24,36–38^

**Figure 1:**
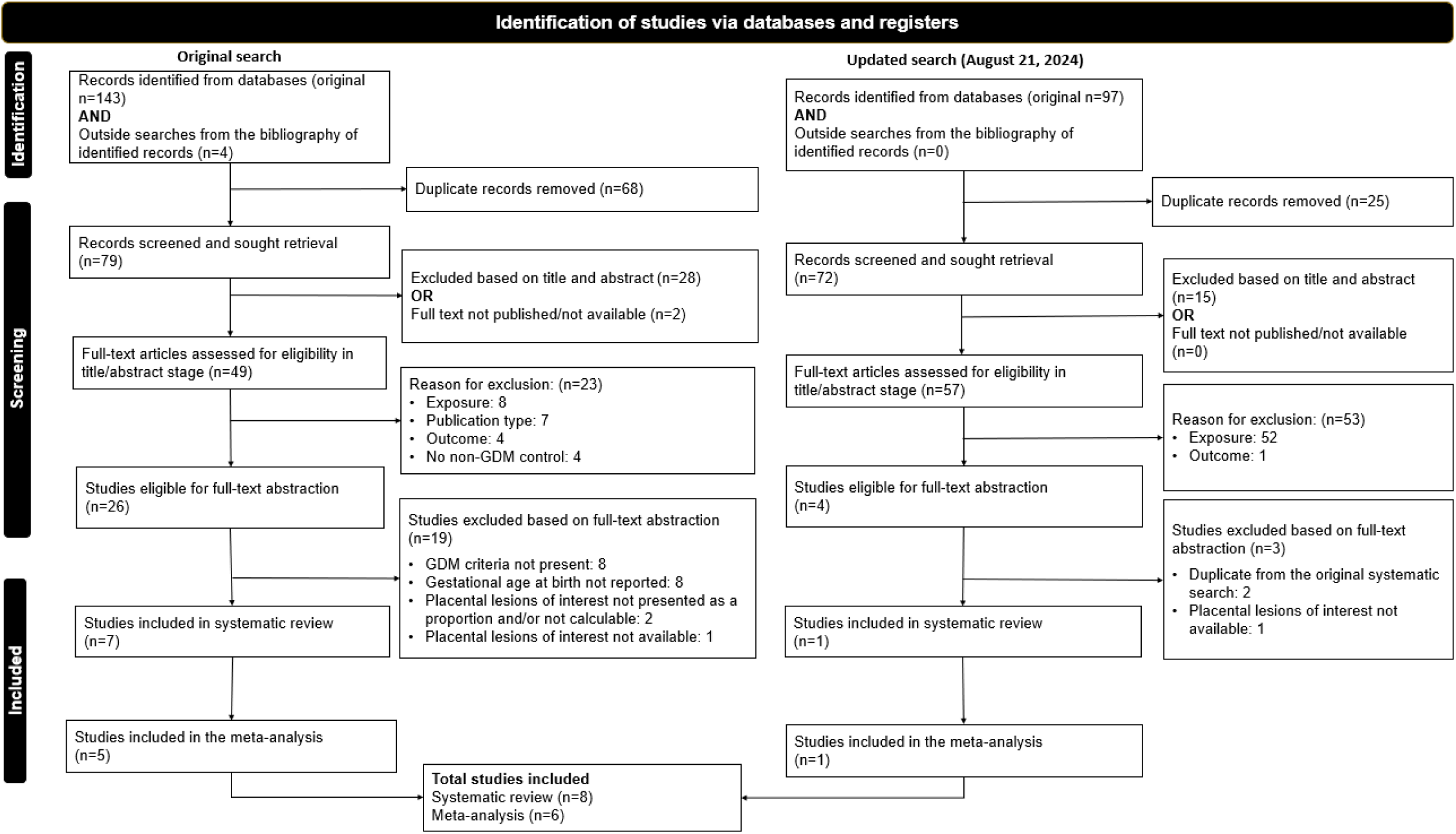
Preferred Reporting Items for Systematic Reviews and Meta-Analyses (PRISMA) study selection.

**Table 1** describes the study design characteristics of all studies included in the systematic review, representing 756 non-GDM pregnancies and 535 GDM pregnancies in eight countries. All studies were case-control, and five GDM criteria were utilized for diagnosis.

**Table 1:**
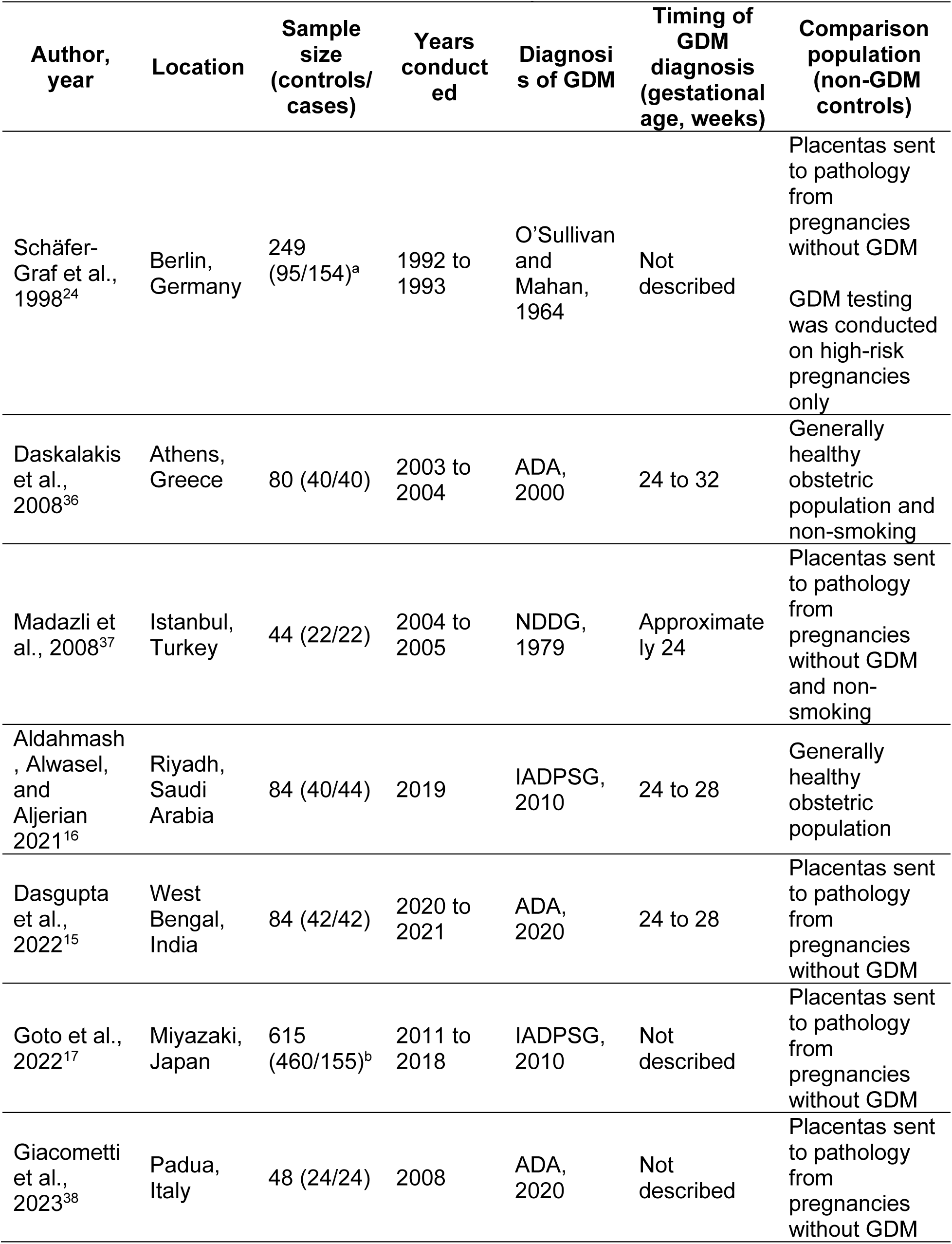
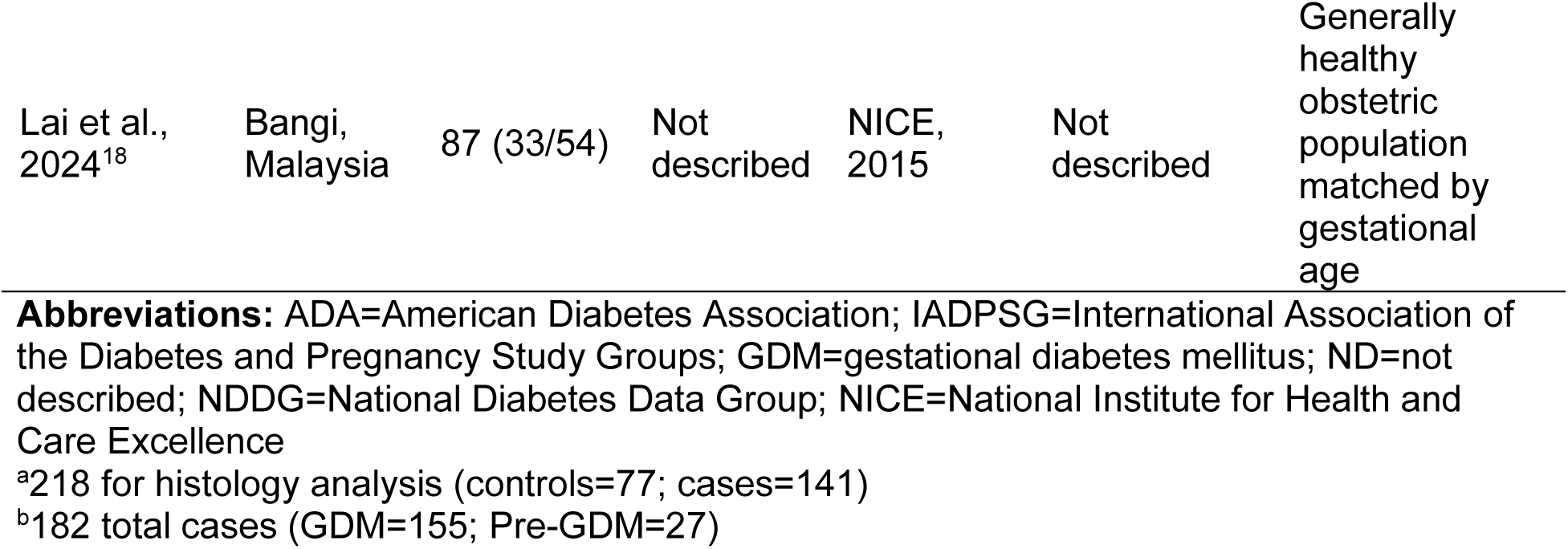
Characteristics of the studies included for systematic review.

Studies were published between 1998 to 2024. Total sample size for non-GDM controls ranged from 22 to 460 and GDM pregnancies ranged from 22 to 155. **Table 2** describes characteristics of the study populations included in the systematic review. The mean (standard deviation (SD)) maternal age in years ranged from 28.2 (5.6) to 33.7 (4.7) in non-GDM controls and 31.2 (5.5) to 34.6 (4.3) in GDM cases. Maternal race and ethnicity, pre-pregnancy BMI, and infant sex (percent male) were not described in most studies. We detailed the studies excluded from the systematic review with reasoning in **Table S3**.

**Table 2:**
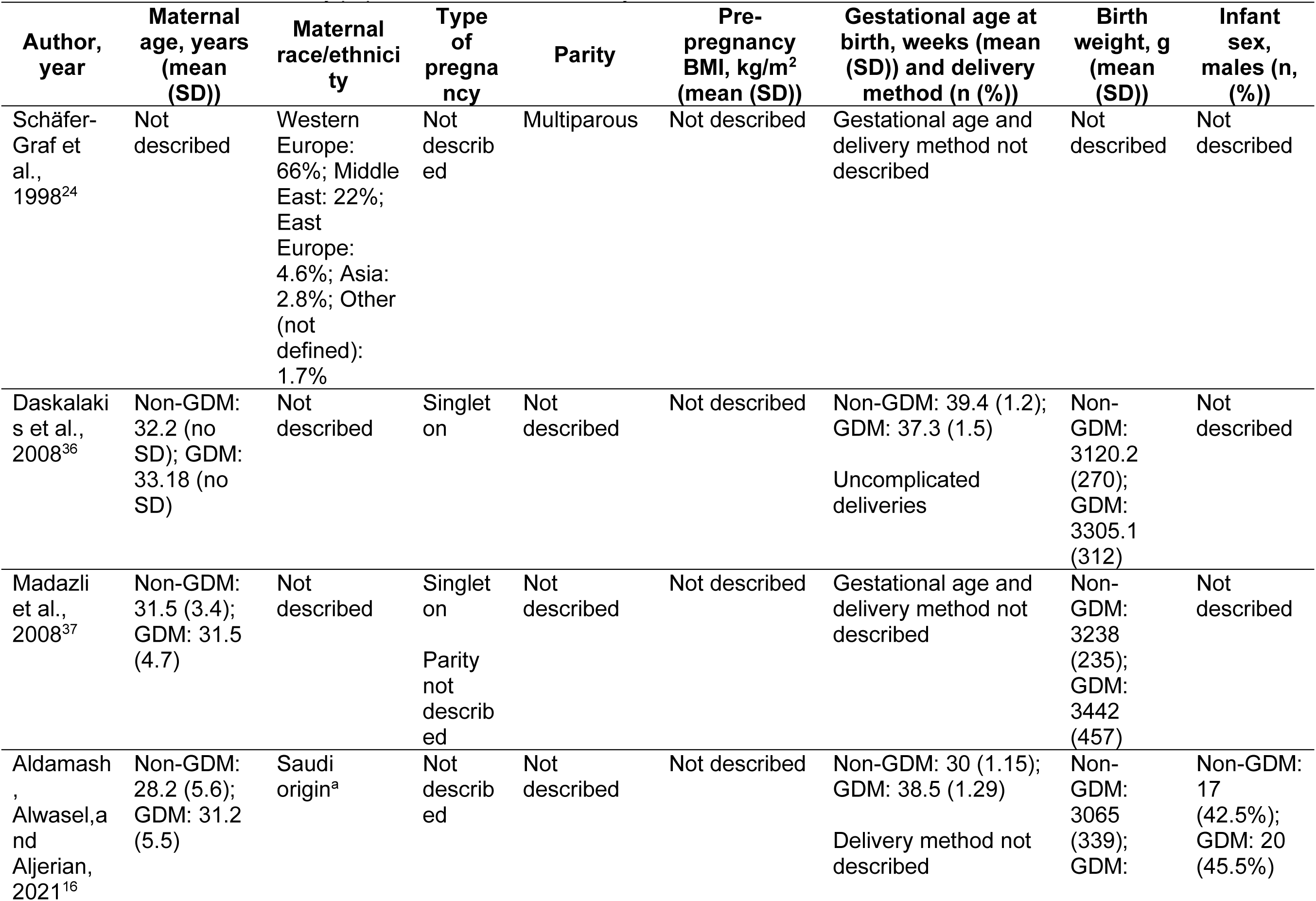
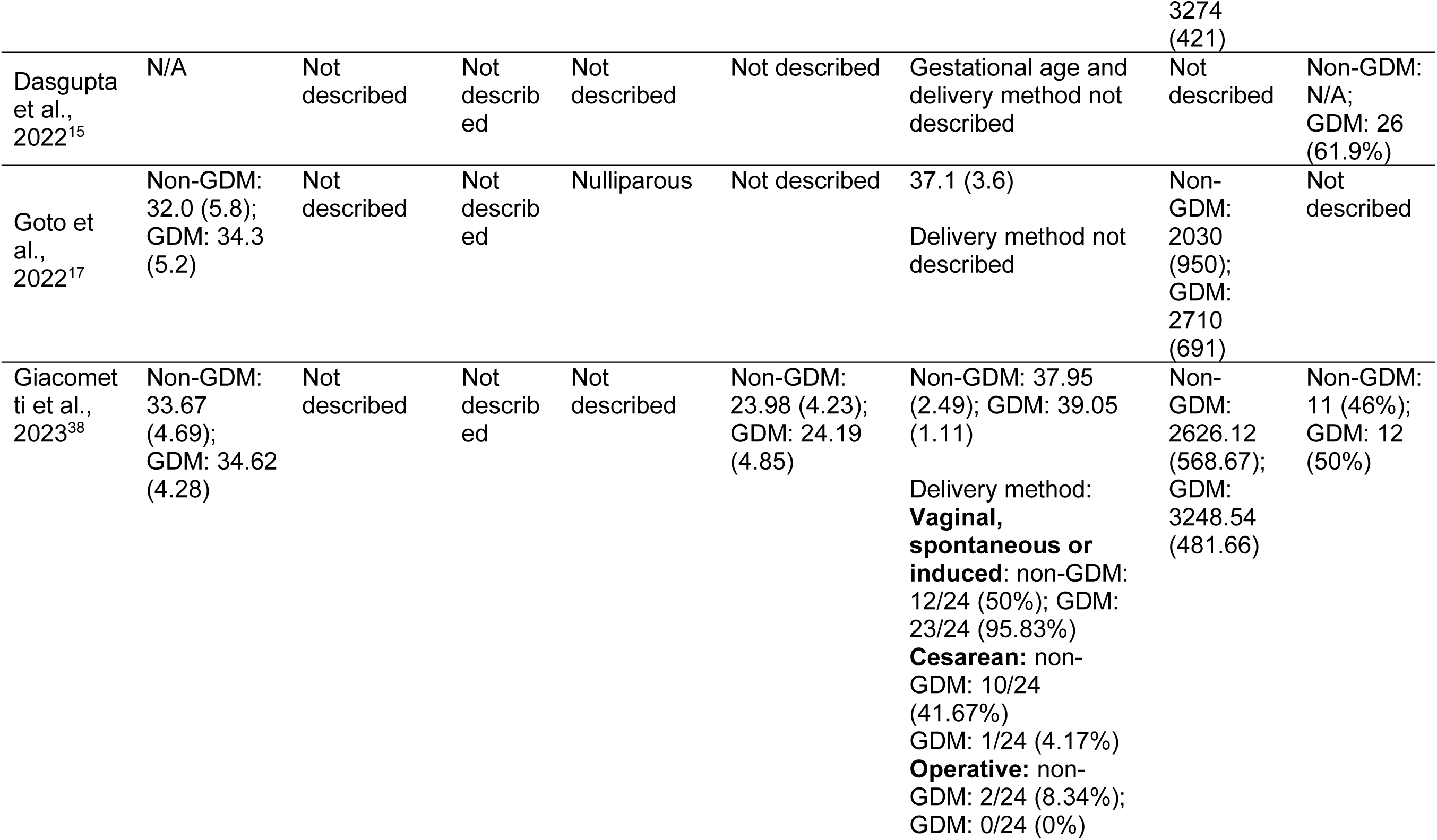
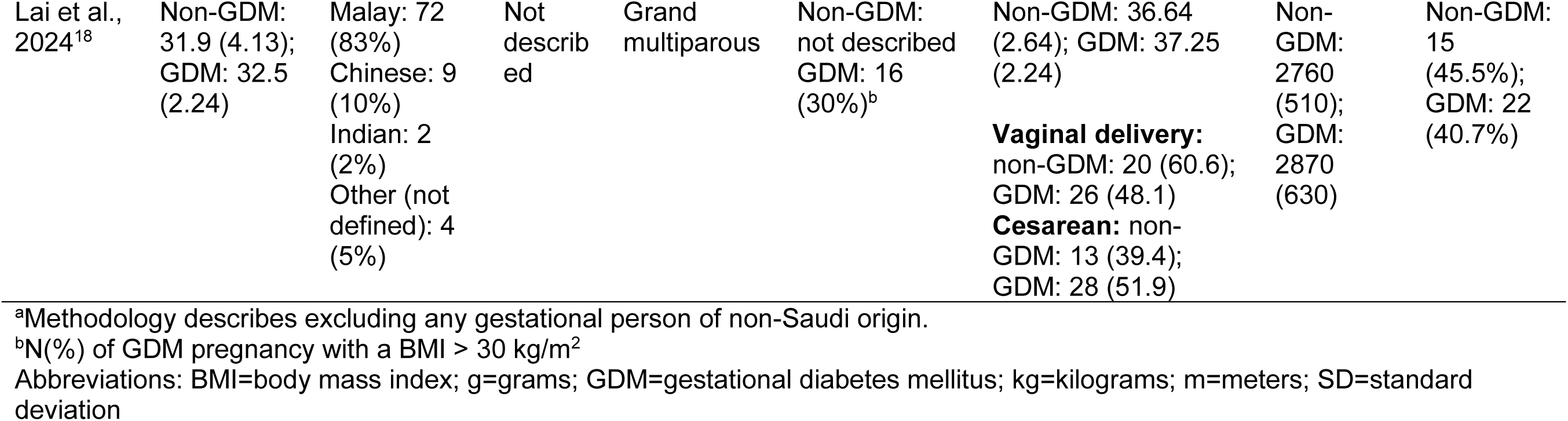
Characteristics of study populations included in the systematic review.

### Risk of bias of included studies

Per the Newcastle-Ottawa Scale, study appraisal scores of all studies eligible for the systematic review are detailed in **Table S4.** The average score was 4.8, with a score range of two to seven. Selection bias was frequently noted. Half of the studies did not completely describe or define their cases to determine if they were representative.^16,24,36,37^ A total of 88% of studies lacked a clear definition of the control population, as the history of GDM or other relevant outcomes in the control group were not explicitly detailed.^16–18,24,36–38^ More than half of the studies were subject to comparability bias related to lack of matching during recruitment or controlling key demographic variables (e.g., age).^15–17,24,36^ Information on the non-response rate was missing from seven studies.^15–17,24,36–38^

### Synthesis of results

We visually summarized the proportional differences in vascular malperfusion lesions in **Figure 2**. Two studies examined AVM,^17,18^ two examined (increased) syncytial knots,^15,16^ six examined DVM,^15,18,24,36–38^ and four examined (increased) fibrin deposition.^15–17^ GDM pregnancies consistently reported greater proportions of DVM, increased syncytial knots, and increased fibrin deposition, when compared to non-GDM pregnancies.

**Figure 2:**
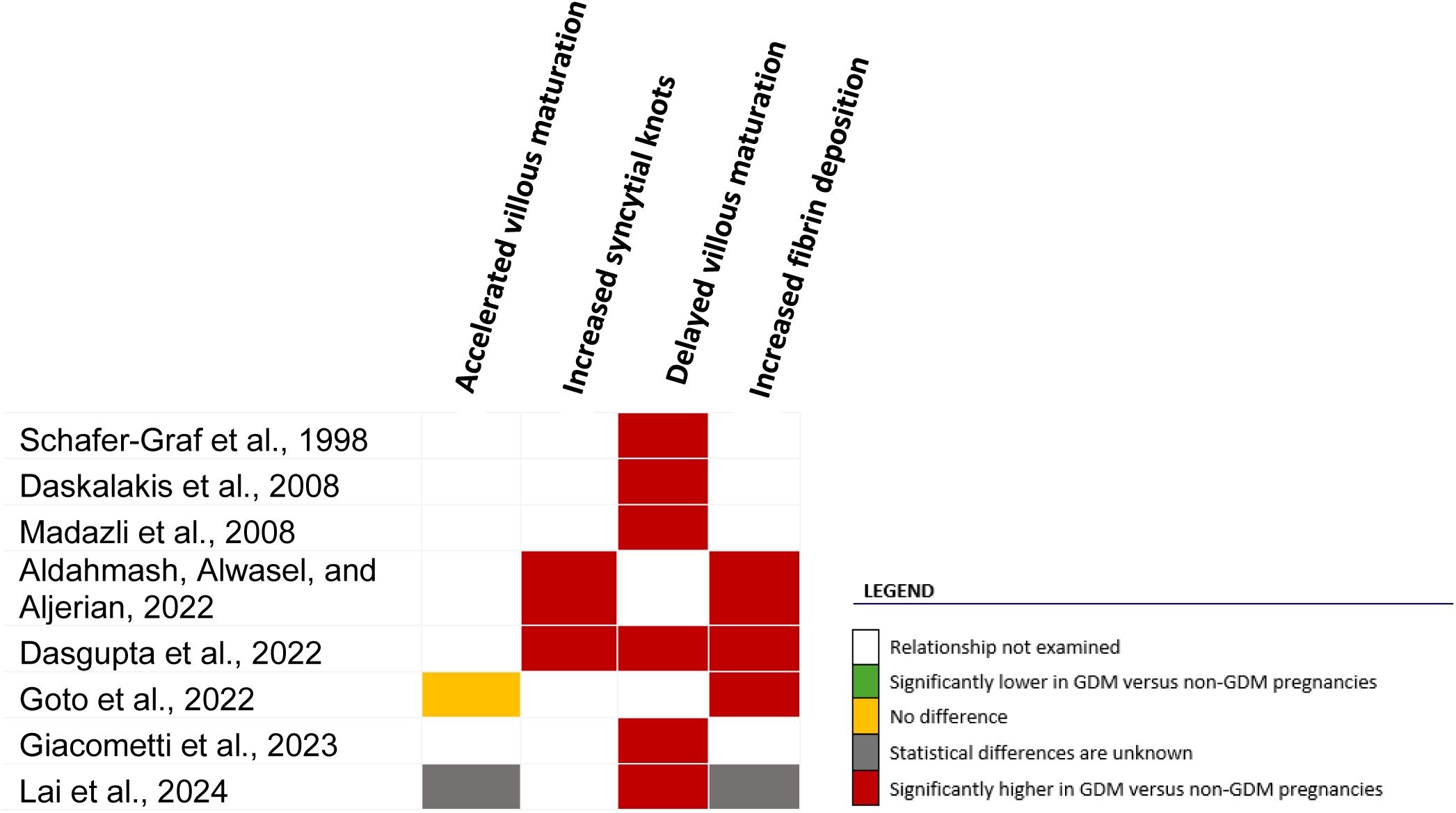
Heatmap of included studies and the proportional differences of vascular malperfusion lesions in GDM pregnancies vs non-GDM pregnancies. Abbreviations: GDM=gestational diabetes mellitus.

We detailed the relevant results of each study in **Table 3**. *Goto et al*. reported that AVM was not different between non-GDM (n=4, 1%) and GDM (n=0, 0%).^17^ *Lai et al.* reported that AVM was lower in the non-GDM controls (n=1, 3%) compared to the GDM cases (n=6, 11%), but statistical differences were not described.^18^ Increased syncytial knots ranged from 19% to 28% in non-GDM controls, and 77% to 86% in the GDM cases.^15,16^ Both studies reported higher proportions of increased syncytial knots in GDM cases compared to non-GDM controls. *Lai et al.* reported syncytial knots as an overall mean and standard deviation but not the proportion of increased syncytial knots by GDM group. *Schäfer-Graf et al*. reported that severe DVM was significantly different between non-GDM controls (n=7/77, 9%) and GDM cases (n=47/141, 33%). All other studies examining DVM had a percent incidence range of 0% to 47.5% in non-GDM controls, and 38% to 83.3% in GDM cases.^15,36–38^ DVM was significantly greater in GDM cases compared to non-GDM controls in most studies.^15,24,36,37^ One study did not test a statistical difference, but a greater proportion of DVM was present in GDM cases compared to the non-GDM controls.^38^ Of note, DVM was combined with maternal vascular malperfusion diagnosis in their non-GDM controls, but not the GDM cases.

**Table 3:**
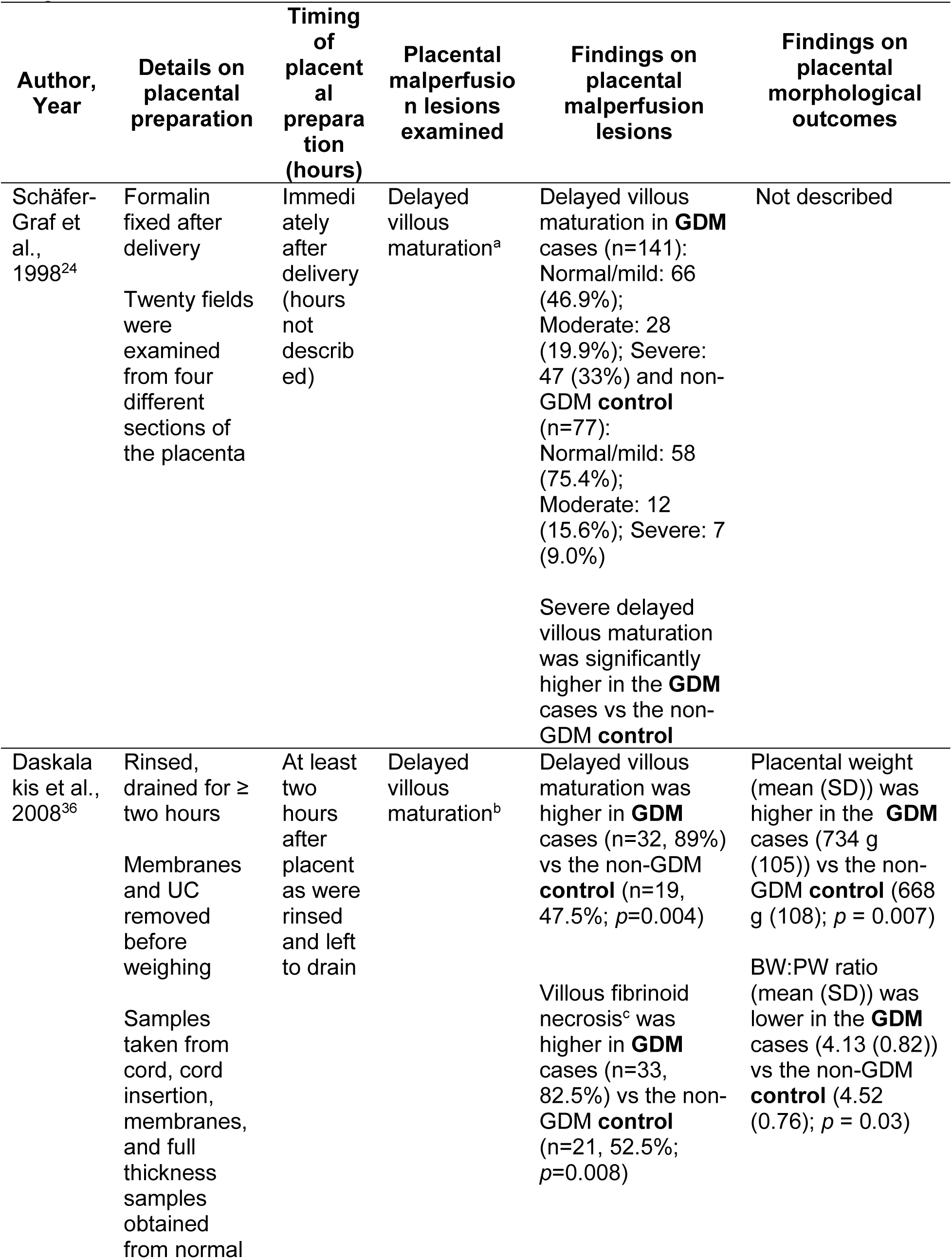
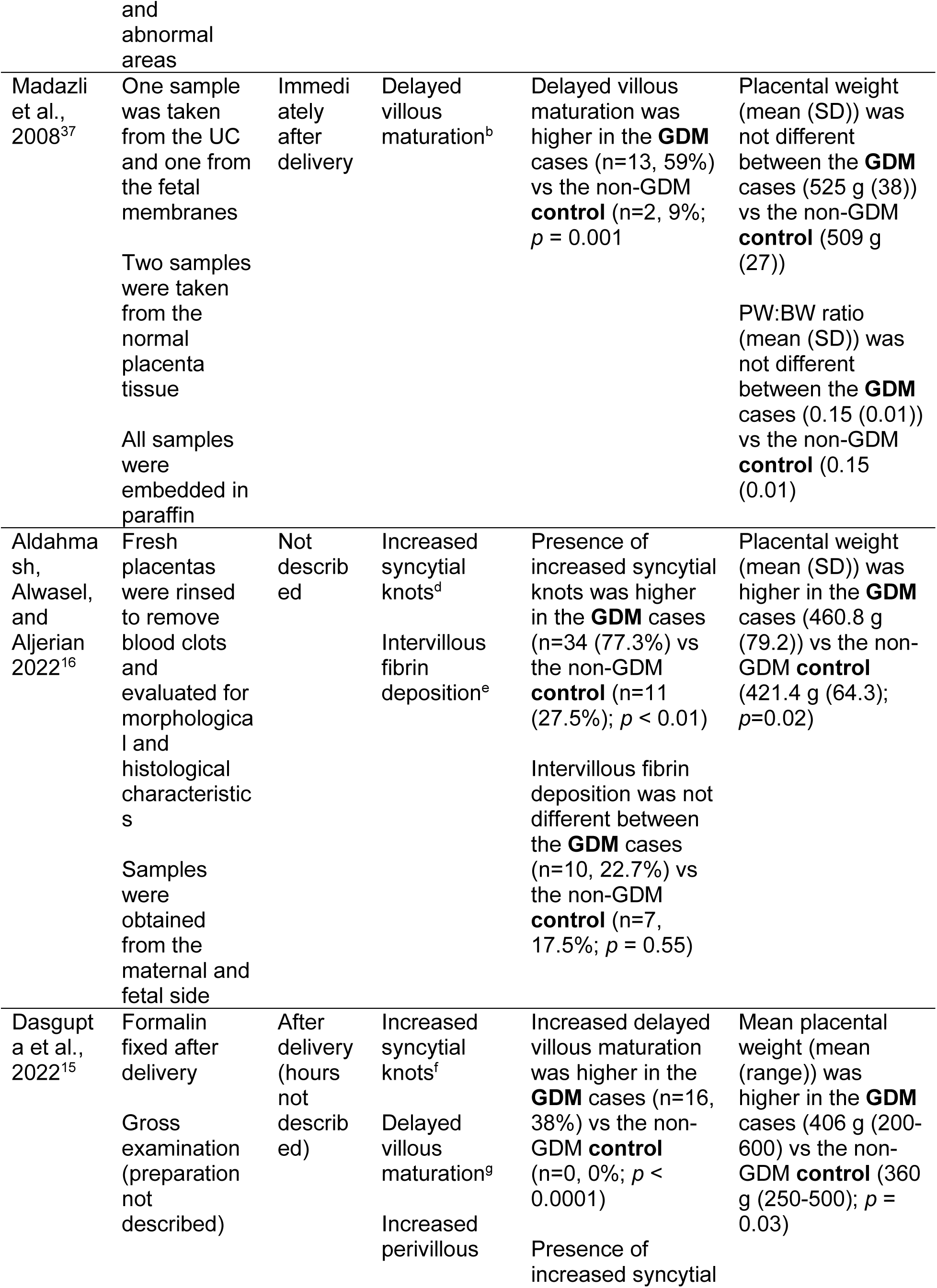
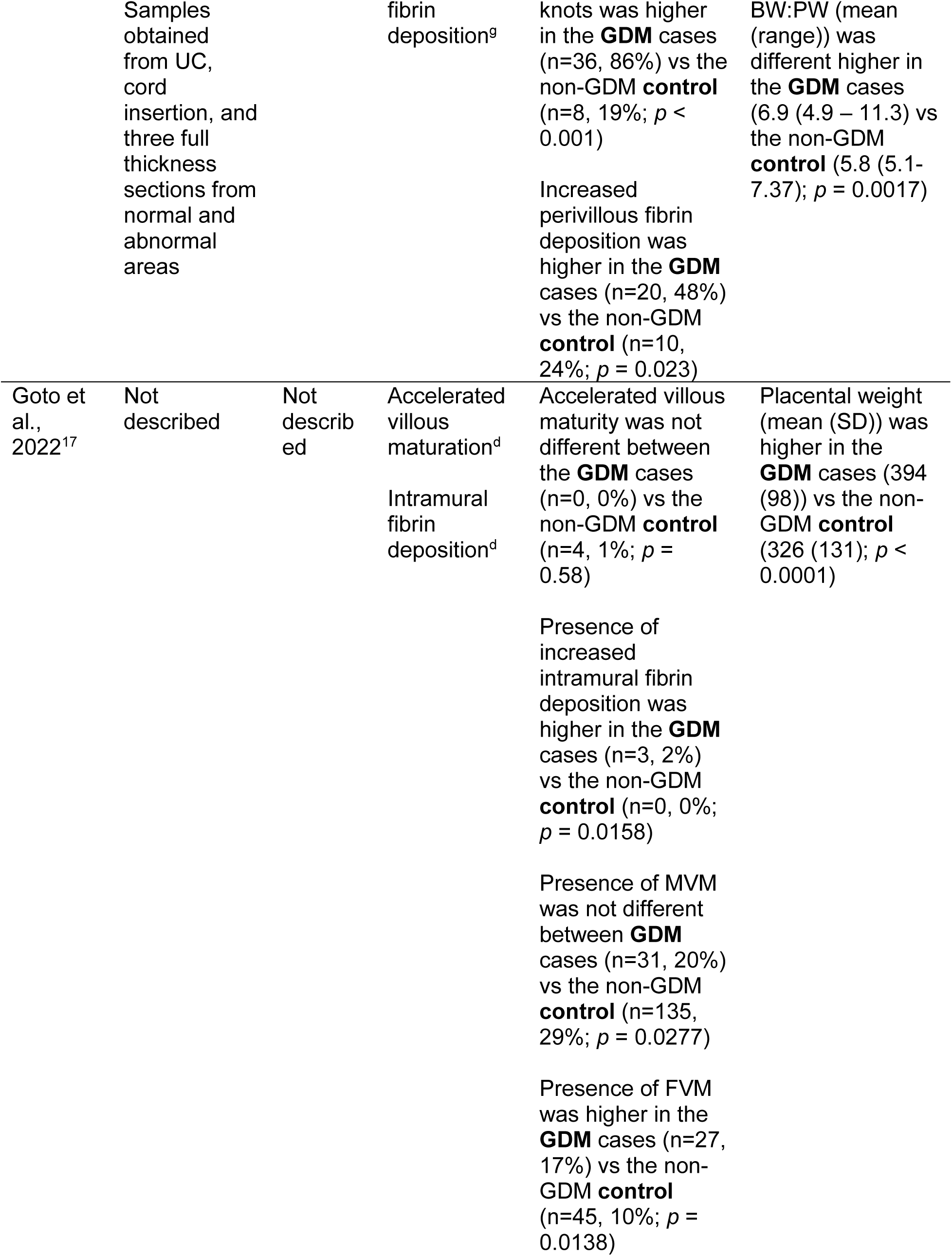
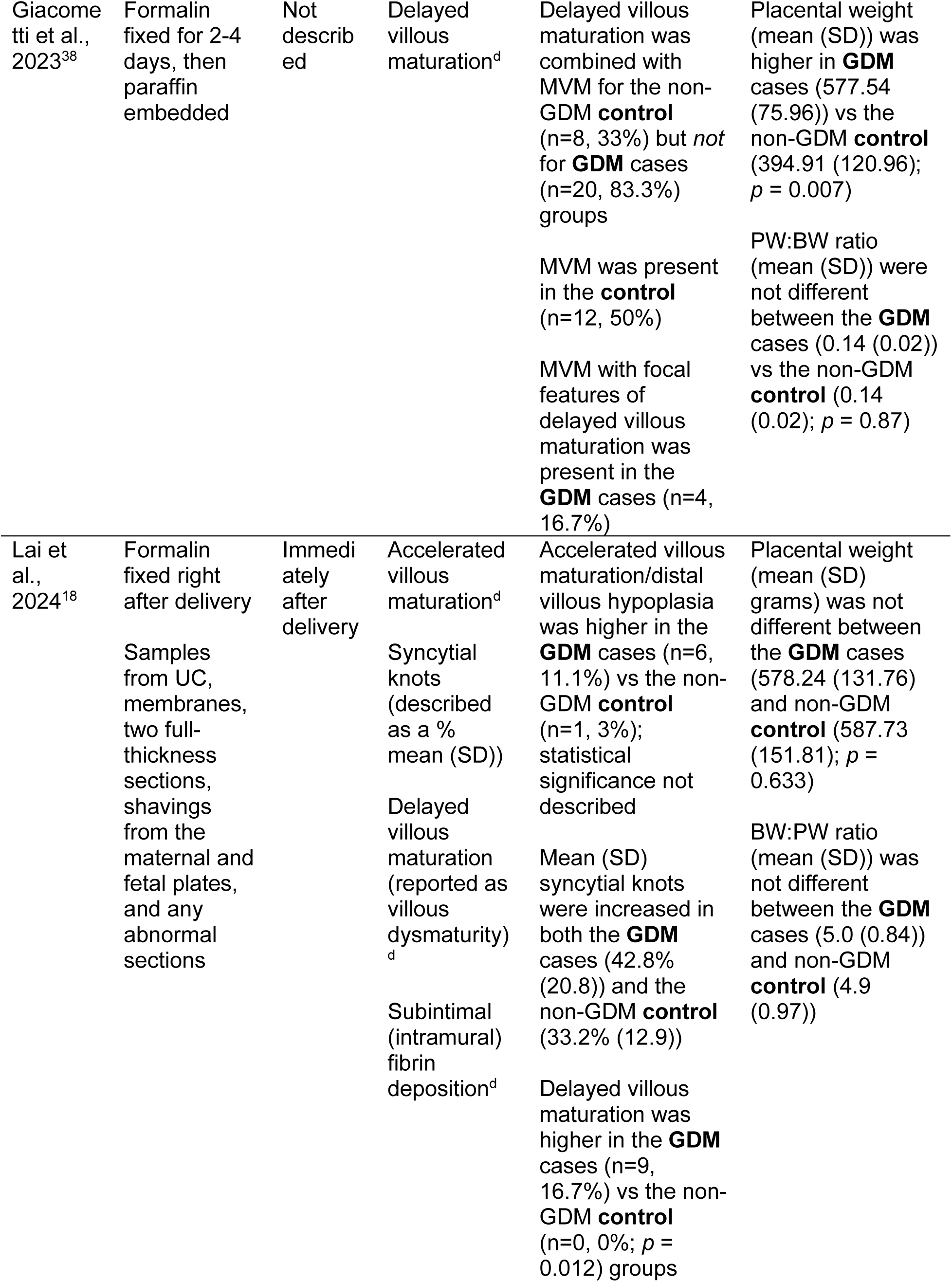
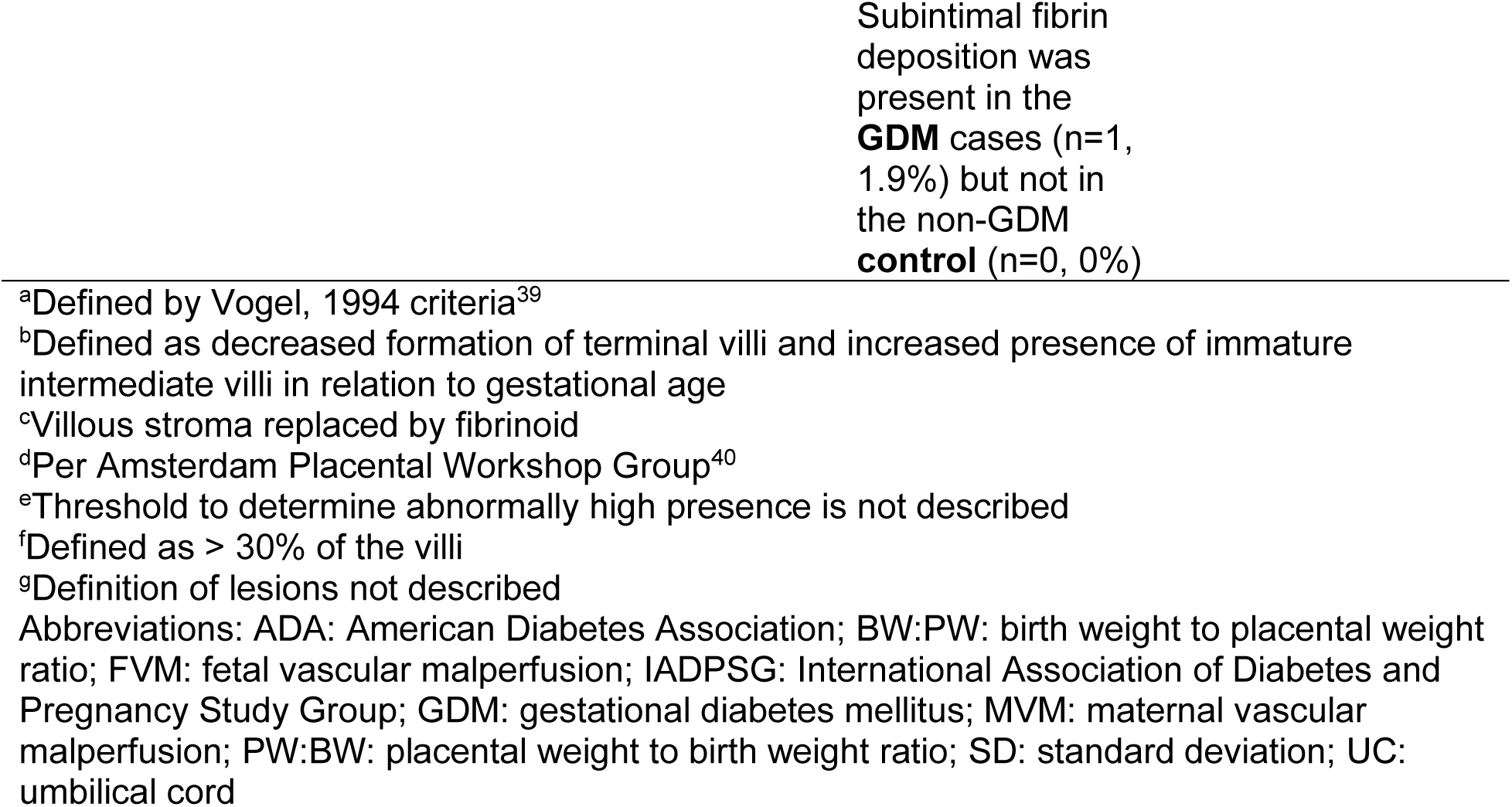
Summary of results for placental malperfusion lesions, placental weight, and the placental/birth weight ratio.

Studies examining increased fibrin deposition sampled from two different locations of the placenta: intervillous/perivillous and intramural.^15–17^ The percent incidence range of fibrin deposition, regardless of location, was 0% to 24% in non-GDM and 2% to 48% in GDM. All three studies reported higher proportions of fibrin deposition, of which two^15,17^ reported significantly higher proportions of fibrin deposition in the GDM cases when compared to the non-GDM controls.

**Figure 3** reports the mixed-effects logistic regression meta-analysis examining GDM and DVM. The unadjusted pooled odds of DVM were six-fold higher in GDM cases compared to non-GDM controls (OR: 6.37 [95% CI: 3.28, 12.37)). Low heterogeneity was present (I^2^ = 29%, τ^2^ = 0.21, *Q* (df=5) = 7.04, *p* value = 0.22). Visual inspection of contour-enhanced funnel plots did not suggest publication bias; however, results should be interpreted cautiously due to the small sample size (n=6 studies; **Figure S1**). *Leave-one-out* sensitivity analysis reported approximately five-to nine-fold higher odds of DVM in GDM pregnancies, compared to non-GDM pregnancies, with influence from *Schäfer-Graf et al. and Daskalakis et al.* (**Figure S2**). Influence analysis confirmed that *Schäfer-Graf et al.* and *Daskalakis et al.* contributed to residual heterogeneity in the model, and both influenced the model fit (**Figure S3**). When these studies were omitted, heterogeneity was lower (I^2^=0%, τ^2^*=*0.00, *Q* (df=2) = 1.07, *p* value = 0.59), and the pooled odds ratio was markedly higher (OR: 13.92 [5.15, 37.67]; **Figure S4**).

**Figure 3:**
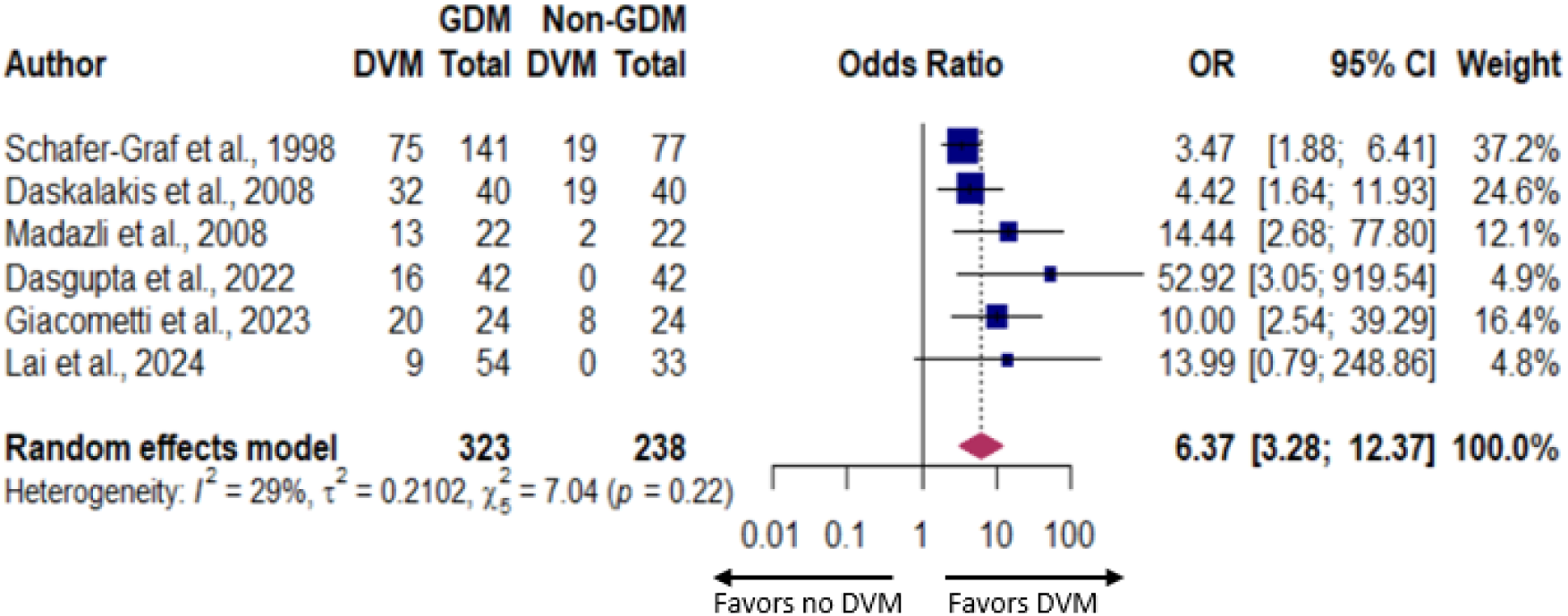
Mixed-effects logistic regression meta-analysis with random study effects examining the association between gestational diabetes mellitus and delayed villous maturation. Abbreviations: CI=confidence interval; DVM=delayed villous maturation; GDM=gestational diabetes mellitus; OR=odds ratio.

## Discussion

In this systematic review and meta-analysis, we found that the literature on GDM and vascular malperfusion lesions was sparse. Our narrative synthesis found that three of four placental lesions were greater in GDM pregnancies compared to non-GDM, and one lesion was not different (in a single study). Meta-analysis determined that exposure to GDM was associated with six-fold greater odds of DVM, compared to pregnancies without GDM.

### Comparison with existing literature

Our systematic search of studies examining AVM and GDM resulted in one study that reported no difference between non-GDM and GDM pregnancies.^17^ The studies determined presence of AVM per Amsterdam Placental Workshop Group criteria, which is a standardized method of diagnosing placental lesions.^17,40^ In AVM, chorionic villi in a preterm placenta resemble a term placenta, with villi fixed by dense syncytial knots, intervillous fibrotic bands, and clusters of distal villi.^14^ There is a distinct pattern alternating between small and sparce to crowded villi. AVM is a signal of maternal vascular malperfusion, which is characterized by derangements to the remodeling of spiral arteries and intervillous blood flow.^12,14^ Past literature does not have a clear consensus on the relationship between AVM and GDM, perhaps due to heterogeneity in study design. *Whittington et al.* found eight-fold greater odds of AVM in pregnancies with diabetes when compared to pregnancies without diabetes.^41^ Different from our objective, in this study GDM was combined with type 1 and type 2 diabetes mellitus, rather than examining GDM alone. Another study by *Siassakos et al.* found that pregnancies with an abnormal glucose value at the GDM screen, but not overt GDM, presented with AVM.^42^ Specifically, they found that 50% (n=6) of diagnosed AVM were in pregnancies with an abnormal glucose reading.^42^ *Scifres et al.* found that 30.5% (n=362) of pregnancies with GDM had maternal vascular malperfusion lesions (which included AVM lesions) and a majority (52.8%) of GDM pregnancies presented with any maternal vascular malperfusion placental lesion, which includes decidual vasculopathy, villous infarction, increased perivillous fibrin deposition, increased intervillous fibrin deposition, and AVM.^43^ However, unlike our study, *Scifres et al*. did not compare these placental lesions to a non-GDM control. The mechanisms of AVM development are not well understood but are thought to be a compensatory response to maternal vascular malperfusion and are diagnosed in preterm placentas.^44^ AVM, and its overarching pathology maternal vascular malperfusion, can result in fetal death, low birthweight, and fetal growth restriction.^11,45,46^ *Christians and Grynspan, 2019* suggested that AVM may be an adaptive response to malperfusion to improve diffusion; thus, it can result in lower odds of fetal death and death from birth to 120 days of age when accounting for gestational age, race, BMI, preeclampsia, placental infection/inflammation, and placental abruption.^47^

Our narrative synthesis found increased syncytial knots in GDM pregnancies, which aligns with past histological analysis.^48–52^ *Aldahmash, Alwasel, and Aljerian (2021)* defined lesions per Amsterdam criteria where syncytial knots are increased if knots are present on more than 33% of villi.^40^ *Dasgupta et al.* reported increased syncytial knots as ≥ 30 knots. Syncytial knots are groupings of syncytial nuclei at terminal villi and is used as a measure of placental maturity.^53^ Syncytial knots are an expected pathology with an average proportion of nearly 30% in a term placenta; however, increased syncytial knots can be a sign of immaturity or malperfusion.^53^ Increased syncytial knots for gestational age are likely a compensatory mechanism in placental formation to maximize transfer of nutrients to the fetus.^53^ *Bhattacharjee et al.* conducted a cross-sectional analysis of placentas sent to pathology.^54^ In contrast to the studies included in our review, they found no significant difference in syncytial knots between normoglycemic controls (n=2 (16.7%)) and GDM pregnancies (n=7 (21.2%), *p*=1.00). However, the same study reported increased syncytial knots in patients with type 1 or type 2 diabetes (n=12 (66.7%), *p* < 0.01) when compared to normoglycemic controls (*p* < 0.01). Clinical intervention was not detailed for any diabetes groups. Even so, increased syncytial knots for gestational age are often present in the GDM placenta and may be a sign of placental malperfusion.

The increased odds of DVM in GDM pregnancies found in our meta-analysis is in line with past literature^50,51,54–56^ and recent evidence suggests an association between DVM and pregnancies with one abnormal glucose value at the GDM diagnostic screen.^42^ Four of the five studies provided definitions for DVM, which varied by criteria, but generally defined DVM as reduced formation of terminal villi, and increased immature intermediate villi for gestational age. DVM is a supportive finding of fetal vascular malperfusion and is characterized by a reduction in the critical vascular branching of the chorionic villi and thus a reduction in vasculosyncytial membrane formation.^10,11,57,58^ *Higgins et al.* conducted a gestational age-matched case-control study and reported significantly higher proportions (n (%)) of DVM in both pre-GDM (criteria for diagnosis not defined; control: 5 (1.8%); pre-GDM: 14 (8%), *p* = 0.02) and GDM (control: 6 (3.4%); GDM: 15 (8.6%), *p* = 0.03).^55^ DVM can result in a host of fetal complications, including fetal death,^11^ making it critical to screen for, and intervene upon, conditions like GDM.

Lastly, our systematic search found an increase in both perivillous/intervillous and intramural fibrin deposition in GDM pregnancies, when compared to non-GDM, albeit in only three studies.^15–17^ Increased fibrin deposition can occur in multiple locations of the placenta. Investigators defined intramural fibrin deposition per Amsterdam Placental Workshop Group criteria.^40^ *Increased intervillous* fibrin deposition was not clearly described by investigators in this review. Perivillous/intervillous fibrin deposition is characterized by excessive fibrin and trophoblasts surrounding the terminal villi.^59^ Intramural fibrin deposition is a different pathology consisting of fibrin bands in the intima of the stem vessels and surface veins of the chorionic plate.^60,61^ *Mayhew and Sampson (2003)* found greater mean (standard error of the mean) fibrin-type fibrinoid in the terminal and intermediate villi of GDM pregnancies (total n =10; 16.7 (2.47)), compared to controls (total n=17, 10.3 (0.77)), and all other diabetes groups.^62^ Importantly, those categorized as GDM included patients with type 2 diabetes, and impaired glucose tolerance. Statistical differences by diabetes group were not conducted, likely in part due to small sample sizes (n < 30 per group). Increased fibrin deposition is a criteria for identifying global partial fetal vascular malperfusion, which is linked to obstruction of the umbilical cord, and by consequence, obstruction to the umbilical vein providing nutrition and oxygen to the fetus.^10^ Strikingly, the presence of increased fibrin deposition can increase the odds of still birth six-fold.^63^

### Strengths and limitations

To our knowledge, this is the first systematic review and meta-analysis to evaluate GDM and lesions from both the maternal and fetal side of the placenta. We conducted a rigorous review of all available literature that was preregistered and followed PRISMA guidelines. Our review has several limitations as well. We established our methods such that all studies were required to include a proportion of lesions and/or a value that was calculable as a proportion. As such, multiple studies presenting histological images showing the presence of lesions in GDM and non-GDM pregnancies were excluded, as they did not have quantifiable results. Our small sample of studies (k=8) limited our ability to run meta-analysis. Only one of our four lesions qualified for meta-analysis, and we were not sufficiently powered to run subgroup analysis.

### Implications and future directions

Our results demonstrated that few published studies have quantified the association between GDM in pregnancy and major lesions of malperfusion in the placenta, highlighting the need for original research in this area. Future systematic reviews should consider examining larger categories such as maternal vascular malperfusion or fetal vascular malperfusion, which encompass multiple lesions associated with GDM. Additionally, subgroup analysis by important characteristics is warranted, including maternal BMI, parity, and gestational age. Studies should consider examining the metabolic milieu that leads to GDM, and its influence on placental malperfusion. Additionally, future studies should examine the relationship between vascular malperfusion lesions and adverse pregnancy outcomes associated with GDM. Chronic and acute conditions are often inextricably linked and may be contributing, with GDM, to derangements in placental vascular development. While more work is needed, this review identified a small body of evidence showing higher proportions of maternal and fetal malperfusion lesions in placentas from pregnancies with GDM.

## Data Availability Statement

The authors confirm that the data supporting the findings of this study are available from the corresponding author (ADG) upon reasonable request.

## Acknowledgments

We wish to extend a special thank you to Dr. Christina Wissinger for her expertise in our systematic search.

## Ethics Approval Statement

Not applicable.

## Author Contributions

All authors contributed to the interpretation and revision of the manuscript. Conceptualization: AA and ADG; Statistical analysis: AA; Writing (original preparation): AA; Writing (reviewing and editing): REW, KG, JAG, ADG. The authors have read and agreed to the preprint version of this paper.

