## Supplementary Materials for "Gestational diabetes mellitus and vascular malperfusion lesions in the placenta: a systematic review and meta-analysis"

**Table S1:** Systematic review search strategy

| **Database** | **Search Terms**[a] |
|---|---|
| PubMed | (("villous"[TIAB]) AND ("immature"[TIAB] OR "immaturely"[TIAB] OR "immatures"[TIAB] OR "immaturities"[TIAB] OR "immaturity"[TIAB])) OR (("delay"[TIAB] OR "delayed"[TIAB] OR "delaying"[TIAB] OR "delays"[TIAB]) AND ("villous"[TIAB]) AND ("maturate"[TIAB] OR "maturated"[TIAB] OR "maturating"[TIAB] OR "maturation"[TIAB] OR "maturational"[TIAB] OR "maturations"[TIAB] OR "mature" [TIAB] OR "matured"[TIAB] OR "matures"[TIAB] OR "maturing"[TIAB] OR "maturities"[TIAB] OR "maturity"[TIAB])) OR (("accelerate"[TIAB] OR "accelerated"[TIAB] OR "accelerates"[TIAB] OR "accelerating"[TIAB] OR "acceleration"[MeSH Terms] OR "acceleration"[TIAB] OR "accelerations"[TIAB] OR "accelerator"[TIAB] OR "accelerator s"[TIAB] OR "accelerators"[TIAB]) AND ("villous"[TIAB]) AND ("maturate"[TIAB] OR "maturated"[TIAB] OR "maturating"[TIAB] OR "maturation"[TIAB] OR "maturational"[TIAB] OR "maturations"[TIAB] OR "maturative"[TIAB] OR "mature"[TIAB] OR "matured"[TIAB] OR "maturer"[TIAB] OR "maturers"[TIAB] OR "matures"[TIAB] OR "maturing"[TIAB] OR "maturities"[TIAB] OR "maturity"[TIAB])) OR (("syncytial"[TIAB]) AND ( “knot” OR "knot s"[TIAB] OR "knots"[TIAB] OR "knotted"[TIAB] OR "knotting"[TIAB]) OR "knot's"[TIAB] OR "knots"[TIAB])) OR ((“fibrinoid” [TIAB] OR “fibrinoids” [TIAB] OR "fibrin"[TIAB] OR "fibrin"[MeSH Terms] OR "fibrins"[TIAB] OR "fibrine"[TIAB]) AND ("deposit"[TIAB] OR "deposit s"[TIAB] OR "deposited"[TIAB] OR "deposites"[TIAB] OR "depositing"[TIAB] OR "deposition"[TIAB] OR "depositional"[TIAB] OR "depositioning"[TIAB] OR "depositions"[TIAB] OR "deposits"[TIAB])) AND (("placenta"[MeSH Terms] OR "placenta"[TIAB] OR "placentas"[TIAB] OR "placenta s"[TIAB] OR "placentae"[TIAB] OR “placental”[TIAB])) AND ("diabetes, gestational"[MeSH Terms] OR "gestational diabetes mellitus"[TIAB] OR "diabetes pregnancy induced"[TIAB] OR "diabetes pregnancy induced"[TIAB] OR "pregnancy-induced diabetes"[TIAB] OR "gestational diabetes"[TIAB] OR "diabetes mellitus gestational"[TIAB] OR “gestational diabetic pregnancies”[TIAB]) |
| BIOSIS and Web of Science | (("villous") AND ("immature" OR "immaturely" OR "immatures” OR "immaturities" OR "immaturity")) OR (("delay" OR "delayed" OR "delaying" OR "delays") AND ("villous") AND ("maturate" OR "maturated" OR "maturating" OR "maturation" OR "maturational" OR "maturations" OR "mature” OR "matured" OR "matures" OR "maturing" OR "maturities" OR "maturity")) OR (("accelerate" OR "accelerated" OR "accelerates" OR "accelerating" OR "acceleration" |

| | |
|---|---|
| | OR "accelerations" OR "accelerator” OR "accelerators") AND ("villous") AND ("maturate" OR "maturated" OR "maturating" OR "maturation" OR "maturational" OR "maturations" OR "maturative" OR "mature" OR "matured" OR "maturer" OR "maturers" OR "matures" OR "maturing" OR "maturities" OR "maturity")) OR (("syncytial") AND (“knot” OR "knots" OR "knotted" OR "knotting")) OR ((“fibrinoid” OR “fibrinoids” OR "fibrin” OR "fibrin" OR "fibrins" OR "fibrine") AND ("deposit" OR "deposits" OR "deposited" OR "deposites" OR "depositing" OR "deposition" OR "depositional" OR "depositioning" OR "depositions" OR "deposits")) AND (("placenta" OR "placentas" OR "placentae" OR “placental”)) AND ("gestational diabetes mellitus" OR "diabetes pregnancy induced" OR "diabetes pregnancy induced" OR "pregnancy-induced diabetes" OR "gestational diabetes" OR "diabetes mellitus gestational" OR “gestational diabetic pregnancies”) |

[a]Filters in addition to search terms: English only, no case reports, no non-human studies, no unpublished studies, only peer-reviewed.

Abbreviations: MeSH: Medical Subject Headings; TIAB: TItle/ABstract

**Table S2:** Diagnostic criteria for Gestational Diabetes Mellitus (GDM)

| Criteria (Year) | Fasting glucose (mg/dL) | Glucose Load (g) | 1-hour glucose (mg/dL) | 2-hour glucose (mg/dL) | 3-hour glucose (mg/dL) |
|---|---|---|---|---|---|
| O'Sullivan and Mahan (1964)[62] | ≥ 90 | 100 | ≥ 165 | ≥ 145 | ≥ 125 |
| NDDG (1979)[63] | ≥ 105 | 100 | ≥ 190 | ≥ 165 | ≥ 145 |
| Carpenter and Coustan (1982)[64] | ≥ 95 | 100 | ≥ 180 | ≥ 155 | ≥ 140 |
| ADA (2000)[65] | ≥ 95 | 100 | ≥ 180 | ≥ 155 | ≥ 140 |
| ADA (2020)[66] | ≥ 95 | 100 | ≥ 180 | ≥ 155 | ≥ 140 |
| WHO (1985)[67] | ≥ 140 | 75 | Not measured | ≥ 200 | Not measured |
| ADIPS (1998)[68] | ≥ 104.4 | 75 | | ≥ 120.6 | |
| WHO (1999)[69] | ≥ 126 | 75 | | ≥ 140 | |
| ADA (2000)[65] | ≥ 95 | 75 | ≥ 180 | ≥ 155 | |
| ADA (2004)[70] | ≥ 95 | 75 | ≥ 180 | ≥ 155 | |
| WHO (2006)[71] | ≥ 126 | 75 | Not measured | ≥ 200 | |
| CDA (2008)[72] | ≥ 95 | 75 | ≥ 191 | ≥ 160 | |
| LADA (2008) | ≥ 100 | 75 | Not measured | ≥ 140 | |
| IADPSG (2010)[73] | ≥ 92 | 75 | ≥ 180 | ≥ 153 | |
| HAPO (2010)[74] | ≥ 92 | 75 | ≥ 180 | ≥ 153 | |
| WHO (2013)[65] | ≥ 92-125 | 75 | ≥ 180 | ≥ 153-199 | |
| ADIPS (2014)[75] | ≥ 92-125 | 75 | ≥ 180 | ≥ 153-199 | |
| NICE (2015)[76] | ≥ 101 | 75 | Not measured | ≥ 140 | |
| ADA (2020)[66] | ≥ 92 | 75 | ≥ 180 | ≥ 153 | |
| Queensland Clinical Guidelines (2021)[77] | ≥ 90 | 75 | ≥ 180 | ≥ 153 | |

Abbreviations: ADA: American Diabetes Association; ADIPS: Australasian Diabetes in Pregnancy Society; CDA: Canadian Diabetes Association; IADPSG: International Association of Diabetes and Pregnancy Study Group; LADA: Latin American Diabetes Association; NDDG: National Diabetes Data Group; NICE: National Institute for Health and Care Excellence; WHO: World Health Organization

**Table S3:** Characteristics of GDM and placental lesions studies that did not meet criteria for inclusion in the systematic review

| Author, year | Country | Years conducted | GDM criteria | Outcome(s) of interest[a] | Reason for exclusion from systematic review |
|---|---|---|---|---|---|
| Verma, Mishra, Kaul, 2010 | India | Not described | Not described | Increased syncytial knots | GDM diagnostic criteria not described<br><br>GA at birth not reported<br><br>Outcomes of interest are not presented as a proportion |
| Pathak et al., 2011 | England | 2007 to 2008 | WHO, 2006 | Fibrin deposition | Control group not clearly described<br><br>GA at birth not reported<br><br>Outcomes of interest are not presented as a proportion |
| Verma, Mishra, Kaul, 2011 | India | Not described | Not described | Increased syncytial knots | GDM diagnostic criteria not described<br><br>GA at birth not reported<br><br>Outcomes of interest are not presented as a proportion |
| Saha et al., 2013 | India | 2006 to 2007 | Non-standardized OGTT criteria | Not present | GA at birth not reported<br><br>Outcomes of interest are not present |

| | | | | | |
|---|---|---|---|---|---|
| Meng et al., 2015 | China | Not described | ADA, 2011 | Increased syncytial knots<br><br>Fibrin deposition | Outcomes of interest are not presented as a proportion |
| Saini et al., 2015 | India | 2012 to 2014 | Not described | Not present | GDM diagnostic criteria not described<br><br>GA at birth not reported<br><br>Outcomes of interest are not present |
| Yavuz et al., 2015 | Turkey | Not described | Non-standardized OGTT criteria | Increased syncytial knots<br><br>Fibrin deposition | GA at birth not reported<br><br>Outcomes of interest are not presented as a proportion |
| Arshad et al., 2016 | Pakistan | 2010 to 2011 | Reports WHO but unclear | Increased syncytial knots<br><br>Delayed villous maturation | GA at birth not reported |
| Bhattacharjee et al., 2017 | India | Not described | Carpenter and Coustan | Increased syncytial knots<br><br>Delayed villous maturation | GA at birth not reported<br><br>Outcomes of interest are not presented as a proportion |
| Ranga et al., 2017 | India | 2016 | Non-standardized OGTT criteria | Not present | Control group were term deliveries only<br><br>GA at birth not reported |

| | | | | | |
|---|---|---|---|---|---|
| | | | | | Outcomes of interest are not present |
| El Sawy et al., 2018 | Saudi Arabia | Not described | IADPSG | Increased syncytial knots | GA at birth not reported<br><br>Outcomes of interest are not presented as a proportion |
| Karunakaran, Nalinakumari, and Ponniraivan, 2018 | India | Not described | Diabetes in Pregnancy Study Group India | Increased syncytial knots | GA at birth not reported<br><br>Outcomes of interest are not presented as a proportion |
| Kamal et al., 2020 | Iraq | 2018 to 2019 | Not described | Increased syncytial knots | GDM diagnostic criteria not reported<br><br>GA at birth not reported<br><br>Outcomes of interest are not presented as a proportion |
| Mohammed and Majid, 2020 | Iraq | 2018 to 2019 | Not described | Increased syncytial knots | GDM criteria not present<br><br>GA at birth not reported<br><br>Outcomes of interest are not presented as a proportion |
| Alqudah et al., 2021 | Ireland | 2014 to 2016 | Not described | Placental maturity<br><br>Increased syncytial knots | GDM diagnostic criteria not reported<br><br>Outcomes of interest are not presented as a proportion |

| | | | | | |
|---|---|---|---|---|---|
| Mehmood et al., 2021 | Pakistan | Not described | Not described | Delayed villous maturation<br><br>Increased syncytial knots | GDM criteria not present<br>GA at birth not reported |
| Nataly et al., 2022 | Israel | 2016 to 2019 | Not described | Not present | Outcomes of interest are not present |
| Whittington et al., 2022 | United States of America | 2007 to 2016 | Carpenter and Coustan | Delayed villous maturation<br><br>Accelerated villous maturation<br><br>Increased syncytial knots | Outcomes of interest are not presented as a proportion |
| Liang et al., 2023 | China | 2018 to 2020 | IADPSG | Delayed villous maturation | Outcomes of interest are not presented as a proportion |
| Luo et al., 2024 | China | 2019 to 2022 | IADPSG | Not present | Outcomes of interest are not present |

Control groups were placentas sent to pathology unless otherwise stated.

[a]One of the following outcomes must be present/calculable as a proportion: delayed villous maturation, accelerated villous maturation, increased fibrin deposition (any kind), and increased syncytial knots

Abbreviations: GDM: Gestational Diabetes Mellitus; IADPSG: International Association of Diabetes and Pregnancy

**Table S4:** Newcastle-Ottawa Scale risk of bias assessment in case-control studies examining gestational diabetes mellitus and placental lesions

| | *SELECTION* | | | | *COMPARABILITY* | *OUTCOME/EXPOSURE* | | | | |
|---|---|---|---|---|---|---|---|---|---|---|
| **Author, Year** | **Is the case definition adequate?** | **Representativeness of cases** | **Selection of controls** | **Definition of controls** | **Comparability of cases and controls on the basis of the design or analysis** | **Ascertainment of exposure** | **Same method of ascertainment for cases and controls** | **Non-response rate** | **Total score (up to 9)** | **Interpretation of score** |
| Schäfer-Graf et al., 1998[26] | * | | | | | * | * | | 3 | Fair |
| Daskalakis et al., 2008[10] | * | | * | | | * | * | | 4 | Fair |
| Madazli et al., 2008[37] | * | | * | | ** | * | * | | 6 | Good |
| Aldahmash, Alwasel, and Aljerian 2022[12] | * | | * | | | * | * | | 4 | Fair |
| Dasgupta et al., 2022[11] | * | * | * | * | | * | * | | 6 | Good |
| Goto et al., 2022[14] | | * | | | | | * | | 2 | Poor |

| | | | | | | | | | | |
|---|---|---|---|---|---|---|---|---|---|---|
| Giacometti et al., 2023[38] | * | * | * | | ** | * | * | | 7 | Good |
| Lai et al., 2024[20] | * | * | * | | * | * | * | * | 6 | Good |

* Denotes whether a study meets the qualifications of the Newcastle-Ottawa Scale assessment

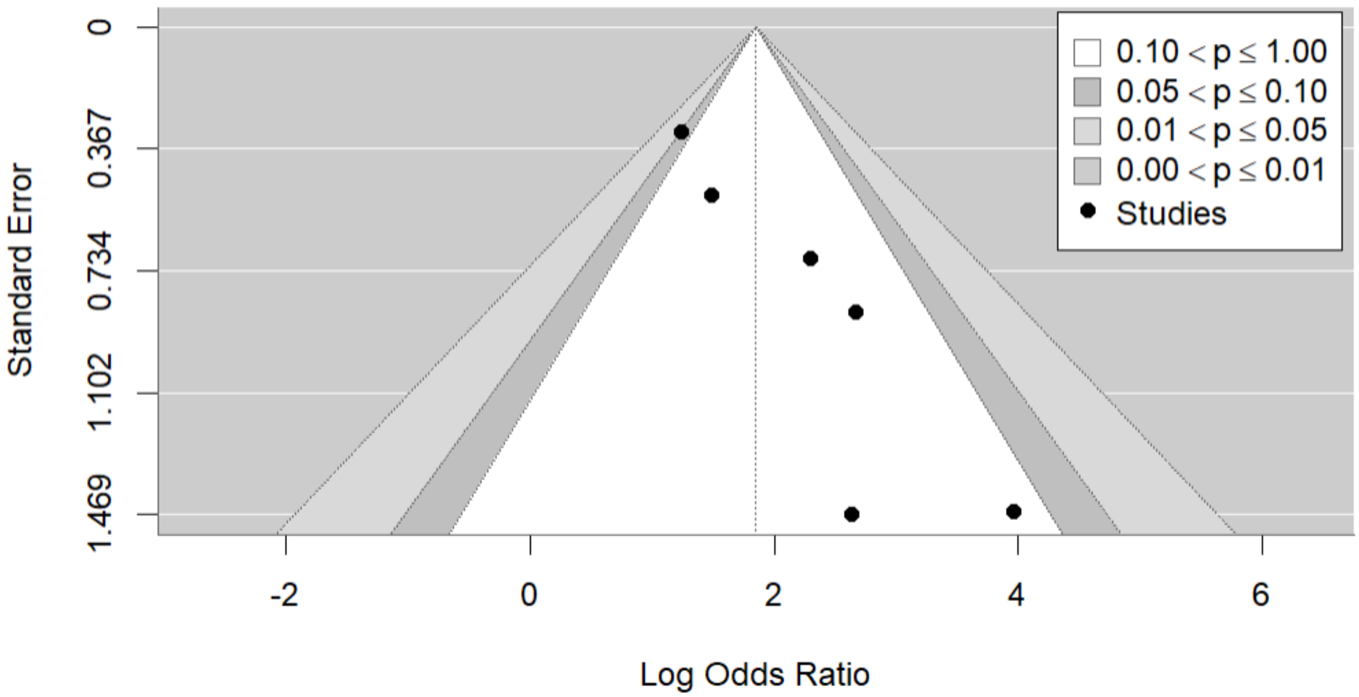


**Figure S1:** Contour-enhanced funnel plot to examine publication bias for studies examining gestational diabetes mellitus and delayed villous maturation.

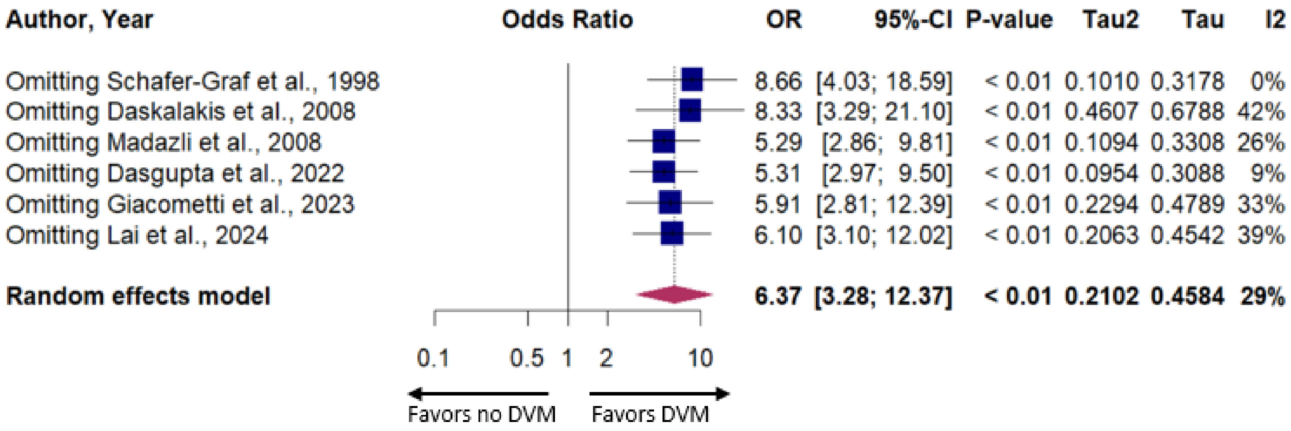


**Figure S2:** Leave-one-out sensitivity analysis examining gestational diabetes mellitus and delayed villous maturation. Abbreviations: CI=confidence interval; OR=odds ratio

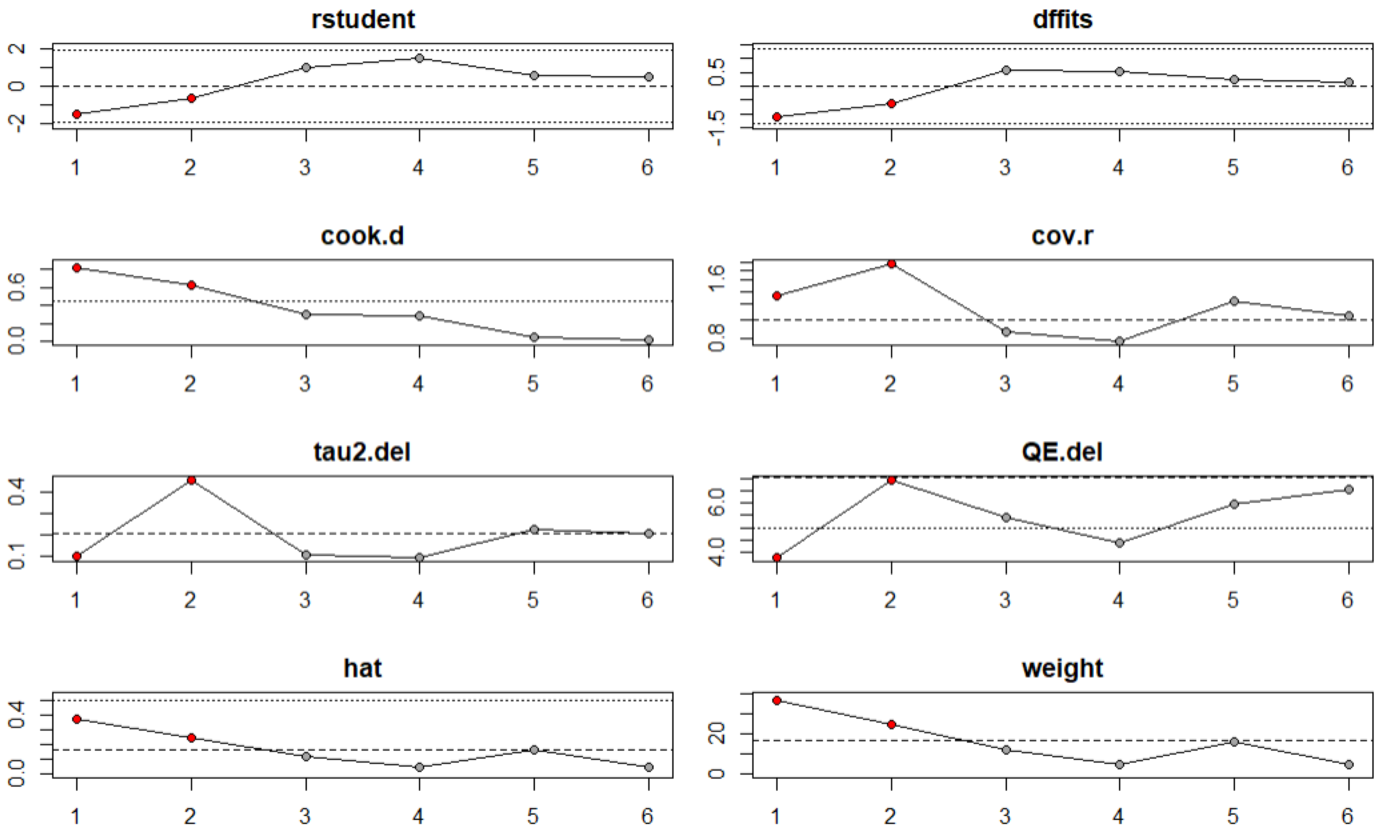


**Figure S3:** Influence analysis of all studies included in the meta-analysis examining gestational diabetes mellitus and delayed villous maturation outcomes. Red circles indicate studies that contribute the most to model fit. Studies are numbered as follows: (1) Schafer-Graf et al., 1998; (2) Daskalakis et al., 2008; (3) Madazli et al., 2008; (4) Dasgupta et al., 2022; (5) Giacometti et al., 2023; (6) Lai et al., 2024. The y-axis values are as follows (left to right, top to bottom): rstudent: studentized residuals, dffits: change in predictions when one study is removed, cook.d: Cook's distance; cov.r: covariance when one study is removed, tau2.del: between-study variance, QE.del: change in Cochran's Q when one study is removed; hat: leverage of each study, weight: weight of each study. Abbreviations: rstudent = studentized residuals; dffits = difference in fits; cook.d = Cook's D; cov.r = covariance ratio; tau2.del = estimate $\tau^2$ when each study is removed; QE.del = test statistic of residual heterogeneity when each study is removed; hat = hat values; weight = weights (%) given to each observed outcome.

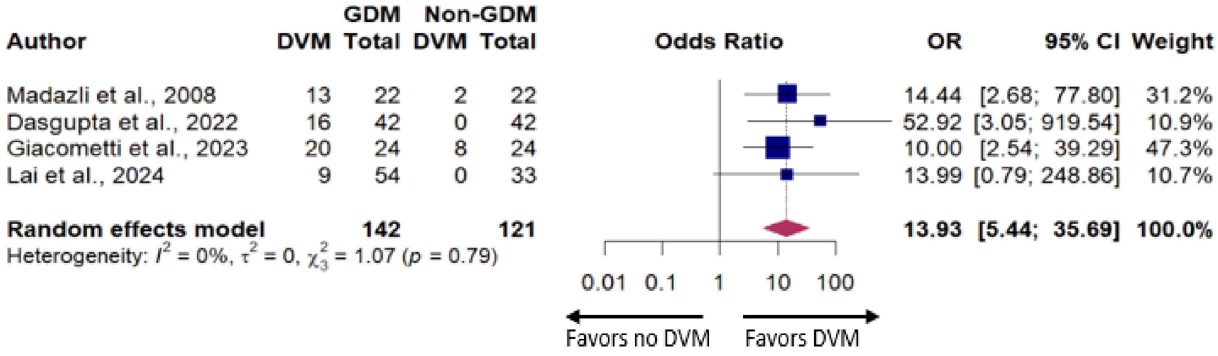


**Figure S4:** Mixed-effects logistic regression model with random study effects after dropping studies (k=2), which contributed the most to the variability in effect size. Abbreviations: DVM=delayed villous maturation; GDM=gestational diabetes mellitus; OR=odds ratio.
